# Variation in psychiatric diagnoses and use of psychotropic medication among children, adolescents, and young adults in Denmark

**DOI:** 10.64898/2026.10.19.26365872

**Authors:** Heidi Sonne, Helene Kildegaard, Mickael Bech, Rikke Wesselhoeft, Anna Sofie Kjærgaard Hansen, Anne Gulbech Ording, Anton Pottegård

**Affiliations:** Clinical Pharmacology, Pharmacy and Environmental Medicine, Department of Public Health, University of Southern Denmark, Campusvej 55, 5230 Odense M, Denmark; Mary Elizabeth’s Hospital, University Hospital Copenhagen, Rigshospitalet, Blegdamsvej 9, 2100 København, Denmark; Department of Political Science and Public Management, University of Southern Denmark, Campusvej 55, 5230 Odense M, Denmark; Research Unit of Child and Adolescent Psychiatry Southern Denmark, Department of Clinical Research, University of Southern Denmark, Campusvej 55, 5230 Odense M, Denmark; Child and Adolescent Psychiatry Southern Denmark, Odense University Hospital, J. B. Winsløws Vej 16, 5000 Odense, Denmark; Mental Health Services Research, Aalborg University Hospital, Mølleparkvej 10, 9000 Aalborg, Denmark; Department of Clinical Medicine, Aalborg University, 9260 Gistrup, Denmark; Danish Center for Health Services Research, Aalborg University Hospital, 9260 Gistrup, Denmark

## Abstract

**Background:** Psychiatric diagnoses and psychotropic medication use among children, adolescents, and young adults have increased in high-income countries, with concerns about geographic variation. We described national trends and geographic variation in Denmark from 2010 to 2025 and developed an interactive online tool to explore these patterns.

**Methods:** We conducted a nationwide, descriptive, repeated cross-sectional register-based study of all Danish residents aged 0-24 years in 2010-2025 (n=2,930,744), using the Danish National Patient Registry and the Danish National Prescription Registry. We estimated annual incidence, prevalence, and, for selected diagnoses, lifetime prevalence, by age and sex, and age- and sex-standardised rates across regions, health councils, and municipalities. We assessed geographic variation using deviations from national means, coefficients of variation, Gini coefficients, and funnel plots.

**Findings:** Psychiatric diagnoses and psychotropic medication use increased from 2010 to 2025. The annual prevalence of attention-deficit hyperactivity disorder (ADHD) increased from 0·41 to 2·3 per 100 individuals, autism spectrum disorders from 0·20 to 1·2, and melatonin use from 0·14 to 3·0. ADHD and autism spectrum disorders were more prevalent among boys and anxiety disorders among girls. Sertraline use was also more prevalent among girls. Geographic variation was observed across diagnoses and psychotropic medications at regional, health council, and municipal levels. For example, lifetime prevalence of autism ranged from 1·3 to 5·9 per 100 across municipalities, and in 2025 regional coefficients of variation ranged from 0·11 (sertraline) to 0·41 (atomoxetine). Results can be explored in the interactive online tool (https://bupstat.sdu.dk/?lang=en).

**Interpretation:** Psychiatric diagnoses and psychotropic medication use increased among Danish children, adolescents, and young adults, with marked geographic variation. Descriptive data cannot establish the causes of this variation, but the findings identify areas for further review and support continued monitoring.

**Funding:** Best for Us project (www.bedstforos.dk), funded by the Novo Nordisk Foundation (NNF24SA0093635) and the Obel Family Foundation (69348).

**Research in context:** *Evidence before this study:* We searched PubMed on 25 September 2026 without date or language restrictions, using two search strings (**Appendix Text 1**). One search string combined search terms for psychiatric disorders and psychotropic medications, children, adolescents, and young adults, and geographic variation. The other search string combined similar population and condition terms with time trends and Denmark or other Nordic countries. We supplemented these with reference lists of key publications, health atlas websites, and national reports. Existing evidence shows substantial increases in psychiatric diagnoses and psychotropic medication use among children and adolescents in high-income countries over the past two decades, with particularly marked rises in attention-deficit hyperactivity disorder (ADHD), autism spectrum disorders, and use of ADHD medications, antidepressants, and melatonin. Studies show considerable geographic variation in diagnostic and prescribing practices that differences in underlying morbidity cannot fully account for. Other countries have produced health atlases examining geographic variation in child and adolescent mental healthcare. However, no comparable nationwide analysis covering both psychiatric diagnoses and psychotropic medication use across multiple geographic levels has previously been conducted in Denmark. The identified studies were predominantly observational and register-based. Nationwide health registries provide high-quality data, but the observational nature limits causal inference about the mechanisms underlying the observed variation.

*Added value of this study:* This study provides the first nationwide, comprehensive mapping of both psychiatric diagnoses and psychotropic medication use among Danish children, adolescents, and young adults over a 16-year period. By applying standardised measures of geographic variation, including coefficients of variation, Lorenz curves, and Gini coefficients, across a broad range of diagnostic categories and medication groups simultaneously, this study extends previous Danish and Nordic work. The accompanying interactive online tool enables dynamic exploration of temporal and geographic patterns and supports ongoing monitoring and dissemination of findings beyond the research community.

*Implications of all the available evidence:* The substantial geographic variation observed across diagnostic and prescribing practices, combined with the rising prevalence of psychiatric diagnoses and psychotropic medication use, highlights the need for continued monitoring of child and adolescent mental healthcare in Denmark. The findings provide a foundation for targeted quality-improvement initiatives, resource planning, and closer alignment of diagnostic and prescribing practice at the regional and municipal levels. Future research should examine the mechanisms underlying geographic variation, including the role of access to services, local clinical cultures, and socioeconomic factors, and evaluate the impact of initiatives to reduce unwarranted variation.

## Introduction

Psychiatric diagnoses and psychotropic medication use among children and adolescents in high-income countries have risen markedly over the past two decades.^1–5^ Geographic variation in diagnostic and prescribing practices has been documented across high-income countries, with differences in population characteristics and healthcare organisation potentially contributing to these patterns.^5–7^ Variation in care that is not justified by patient needs or preferences is considered unwarranted and may arise from multiple factors, including inconsistent clinical practices, differences in access to specialist care, supplier-induced demand, or local referral or treatment cultures.^8^ While some variation is expected given differences in population needs and demographic composition, large variation across regions or municipalities raises concerns about adherence to evidence-based guidelines, equitable access to care, and overall treatment quality.^8^

Describing these geographic patterns is an important first step towards identifying unwarranted variation. Such evidence can inform planning and quality improvement efforts to ensure appropriate, evidence-based care. Health atlases, which systematically map geographic variation in healthcare use and outcomes across administrative areas, represent one such approach. Such atlases have been developed for child and adolescent mental healthcare in several countries,^9,10^ and evidence from other healthcare settings shows that atlas findings can contribute to developing clinical standards and system-level initiatives to improve care.^11,12^ However, no nationwide publicly accessible health atlas has systematically presented temporal and geographic variation in both psychiatric diagnoses and psychotropic medication use among Danish children and adolescents.

In this study, we aimed to describe temporal and geographic variation in psychiatric diagnoses and psychotropic medication use among children, adolescents, and young adults in Denmark from 2010 to 2025. We used standardised metrics across regions, health councils, and municipalities to identify potential unwarranted variation that may serve as targets for quality improvement. We also developed an interactive online atlas to help explore these patterns and demonstrate a population-based monitoring approach that may be transferable to other registry-based healthcare systems.

## Methods

This nationwide, descriptive, repeated cross-sectional study examined temporal and geographic variation in psychiatric diagnoses and psychotropic medication use among Danish children, adolescents, and young adults aged 0–24 years from January 1^st^, 2010, to December 31^st^, 2025. We also developed an interactive online visualisation tool to help users explore these patterns. This study is reported according to the STROBE statement and its extension for routinely collected health data (RECORD).^13,14^

### Setting

Denmark has a universal, tax-funded healthcare system. It provides diagnostic assessments, consultations, and hospital-based services free of charge, whereas outpatient prescription medications are subject to a graduated copayment system based on annual medication expenditure.^15^ Denmark’s healthcare system is organised into five regions, which are responsible for hospitals, general practitioners, and specialists in private practice under regional contracts.^15^ The regions are subdivided into 98 municipalities, which are grouped into 17 health councils. With universal, tax-funded healthcare and nationwide registers covering the entire population, Denmark offers a setting where geographic differences are less likely to reflect insurance coverage or out-of-pocket costs than in many other health systems.

Child and adolescent mental health services in Denmark are primarily organised as public specialised regional services. Children and adolescents are generally treated in child and adolescent mental health services until age 18, at which point care is transferred to adult psychiatric services, although some regions allow continued follow-up beyond this age.^16^ Psychiatric diagnoses in individuals under age 18 years are predominantly assigned within child and adolescent mental health services. Assessment and treatment may also be provided in other settings, including paediatric departments and psychiatrists in private practice, with or without public reimbursement agreements.^17^ Non-pharmacological interventions are generally recommended as first-line treatment, and initiation of psychotropic medication in children and adolescents is recommended to be managed by specialists in child and adolescent psychiatry.^17^

### Data sources

We obtained individual-level data from multiple Danish national registers and linked them using the unique personal identification number assigned to all individuals living in Denmark.^18^ Individual-level data were accessed through the secure server at the Danish Health Data Authority.

The Danish Civil Registration System contains information on all residents of Denmark since 1968, including birth date, sex assigned at birth, date of death, and place of residence.^18^

The Danish National Patient Registry holds information on all somatic and psychiatric inpatient and outpatient hospital contacts since 1995, including the date of contact and diagnoses according to the International Classification of Diseases, 10^th^ edition (ICD-10).^19,20^ The Danish National Patient Registry has been validated for a range of psychiatric diagnoses.^20^

The Danish National Prescription Registry contains individual-level data on all prescriptions filled at Danish pharmacies since 1995, including dispensing dates and indications for use.^21^ The registry does not include in-hospital or over-the-counter medication use, and all medications listed are classified according to the Anatomical Therapeutic Chemical (ATC) classification system.^22^ The Danish National Prescription Registry has high completeness for redeemed prescriptions.^21^

The Registry of Health Care Providers contains information on all healthcare professionals authorised to practice in Denmark, including their professional credentials, workplace affiliations, and specialisations.^23^ Prescribers were classified by prescriber setting (private or hospital) and by speciality code. For prescribers in private practice, speciality was defined by the most recently registered main speciality. For hospital prescribers, speciality was determined from department codes and thus reflected the issuing department rather than the individual physician. Prescriptions without an identified prescriber or speciality were classified as unknown.

### Study population

The study population consisted of all Danish residents aged 0–24 years between 2010 and 2025. Individuals were categorised by sex assigned at birth and by age on July 1^st^ each year. We defined four age groups (0–5, 6–11, 12–17, and 18–24 years) and two aggregate groups (0–17 and 0–24 years). We used a full-population sample and excluded no individuals from the national prevalence analyses. Incidence analyses were restricted as described below.

### Diagnoses and medications

We identified psychiatric disorder diagnoses using ICD-10 codes recorded in the Danish National Patient Registry.^19,20^ The following diagnostic categories were included: schizophrenia spectrum disorder (F20–29), bipolar disorder (F30–31), depressive episode (F32–33), anxiety disorders including obsessive-compulsive disorder (F40–48 plus F93), eating disorders (F50), personality disorders (F60–69), intellectual disability (F70–79), autism spectrum disorders (F84.x excluding F84.2–F84.4), attention-deficit hyperactivity disorder (ADHD) and attention-deficit disorder (ADD) (F90 and F98.8), attachment disorders (F94.x excluding F94.0) and tic disorders (F95) (**Appendix Table 1**). These categories were inspired by the spectrum of mental disorders in children and adolescents as defined in a Danish cohort study.^24^ Diagnoses were included irrespective of the hospital department in which they were recorded (psychiatric or somatic, including paediatric departments).

Psychotropic medication use was identified using ATC codes from the Danish National Prescription Registry.^21^ Medications spanned antipsychotics, anxiolytics, and hypnotics/sedatives (ATC N05) and antidepressants and ADHD medications (ATC N06). Specific ATC codes for the individual medications mentioned are listed in **Appendix Table 2**.

### Statistical analyses

We estimated annual incidence and prevalence proportions per 100 individuals for each psychiatric diagnosis and psychotropic medication from 2010 to 2025. The study covers the entire population, and we therefore report proportions without confidence intervals. Nationwide registers with prospectively recorded data, consistent definitions across years and geographic areas, and standardisation for age and sex limit bias.

We defined prevalence as at least one hospital contact with a recorded diagnosis or at least one prescription redemption within a given calendar year. For selected diagnoses with a typically long-term course, such as autism spectrum disorder and ADHD, lifetime prevalence was also estimated, defined as any prior diagnosis in the current year or before since 1995 (**Appendix Table 1**). We defined incidence as the first recorded diagnosis or prescription redemption, with no earlier record of the relevant diagnosis or medication since 1995. For psychotropic medications, analyses were conducted across all four hierarchical levels of the ATC classification system (from broad therapeutic groups to specific substances), and individuals were counted once per category in each year but could contribute to several categories.

In the prevalence analyses, the denominator included all individuals alive and resident on July 1^st^ of each year within the relevant age range and sex. In the incidence analyses, we restricted the numerator and denominator to individuals with continuous observation for at least the preceding five years, or since birth for individuals younger than five years. We defined continuous observation as uninterrupted residence in Denmark. The denominator thus comprised individuals who met this requirement on July 1st of each year and had no record of the relevant diagnosis or medication since 1995.

Geographic analyses included only individuals with a known residential address (>98% each year), while individuals with a missing address remained in the national analyses. We masked estimates based on fewer than five individuals in the numerator or denominator to protect confidentiality.

At the national level, we calculated incidence and prevalence, as well as the distribution of recorded indications for each medication group, and prescriber types (general practitioners, private psychiatrists, other private practitioners, hospital psychiatrists, other hospital practitioners, and unknown) (**Appendix Table 3**). At the regional, health council, and municipal levels, we estimated age- and sex-standardised incidence and prevalence using the national age- and sex-distribution. We quantified geographic variation in different ways: as the percentage deviation of each area from the national mean; as the coefficient of variation (the standard deviation of the area-specific estimates divided by the national estimate); and, at the municipal level, using inequality measures based on Lorenz curves and corresponding Gini coefficients. Lorenz curves describe the cumulative share of incidence or prevalence, summed across geographic units, with the units ranked by rate. The Gini coefficient summarises inequality on a scale from 0 (no inequality) to 1 (maximum inequality). To further assess geographic variation and distinguish systematic variation from random fluctuation, we constructed municipal-level funnel plots. In these plots, we plotted municipality-specific incidence or prevalence estimates against the number of inhabitants in the relevant age group and sex in each municipality, obtained from Statistics Denmark. Control limits were set at 95% and 99·8% and represent the expected range of random variation around the national mean under a binomial assumption.

Analyses were conducted using R version 4.4.3 and were validated internally by a data manager not otherwise involved in the study.

### Online visualisation tool

We developed an interactive online visualisation tool (https://bupstat.sdu.dk/?lang=en) to provide dynamic access to the study results using R Shiny. The tool is based on the results from the statistical analyses and presents estimates of incidence, prevalence, lifetime prevalence (where applicable), and geographic variation. The tool enables interactive visualisation of annual trends from 2010 to 2025, stratified by age group and sex, and allows examination of patterns at national, regional, health council, and municipal levels. Users can additionally explore prescribing setting and recorded indications for psychotropic medications.

### Ethics and approvals

The study was registered at the University of Southern Denmark (Research and Innovation Organisation, no. 11.106). According to Danish law, studies based solely on register data do not require ethics approval or informed consent.

## Results

The study included 2,930,744 unique children, adolescents, and young adults aged 0–24 years residing in Denmark between 2010 and 2025. The annual number of individuals ranged from 1,651,616 to 1,689,592 between 2010 and 2025. The sex distribution was balanced, with 51% boys and 49% girls in both 2010 and 2025 (**Supplementary Table 1**). Complete results for all psychiatric diagnoses and psychotropic medications are available in the online tool (https://bupstat.sdu.dk/?lang=en).

Here, we present results for the most prevalent psychiatric diagnoses and psychotropic medications, as well as selected examples with particularly high geographic variation. These examples illustrate the scope of the analyses and highlight key findings.

### Time trends

The annual recorded prevalence increased for all of the most prevalent psychiatric diagnoses among individuals aged 0–24 years between 2010 and 2025, most notably for ADHD (from 0·41 to 2·3 per 100 individuals), autism spectrum disorders (from 0·20 to 1·2), and anxiety disorders (from 0·52 to 1·3) (**Figure 1**). In 2025, the prevalence was 0·35 per 100 for depression, 0·34 for schizophrenia, 0·21 for personality disorders, and 0·21 for intellectual disability, compared with 0·24, 0·15, 0·17, and 0·12 in 2010.

**Figure 1.**
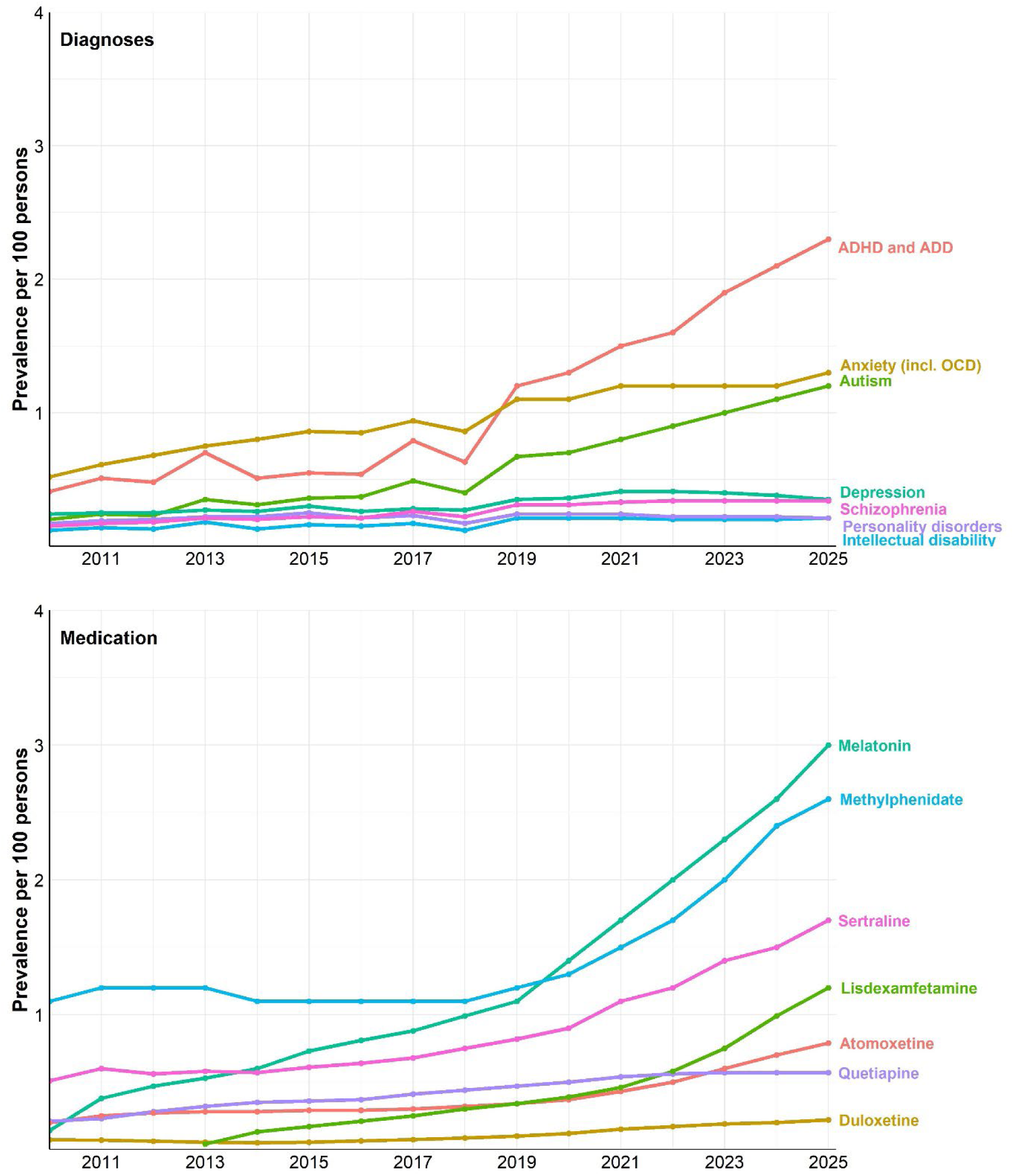
Annual prevalence of the most common psychiatric diagnoses and psychotropic medications among Danish residents aged 0–24 years, 2010–2025. Annual prevalence per 100 persons for selected psychiatric diagnoses and psychotropic medications among Danish residents aged 0–24 years from 2010 to 2025. Estimates are based on individuals alive and residing in Denmark on July 1^st^ of each year. ADD: attention-deficit disorder; ADHD: attention-deficit hyperactivity disorder; OCD: obsessive-compulsive disorder.

The prevalence of psychotropic medication increased, particularly for melatonin (from 0·14 to 3·0 per 100), methylphenidate (from 1·1 to 2·6), sertraline (from 0·51 to 1·7), and lisdexamfetamine (from 0·04 in 2013, to 1·2) (**Figure 1**). In 2025, the prevalence was 0·79 per 100 for atomoxetine, 0·57 for quetiapine, and 0·22 for duloxetine, compared with 0·20, 0·21, and 0·07 in 2010.

### Sex and age variation

Sex differences are presented for the most prevalent psychiatric diagnoses and psychotropic medications (**Figure 2**). Autism and ADHD diagnoses were more prevalent among boys, whereas anxiety diagnoses were more common among girls. Sertraline use was also more prevalent among girls. Melatonin use was slightly higher among boys until 2020 and among girls from 2021 onward. The difference in the level of methylphenidate use between boys and girls decreased over time, and in 2025 the prevalence was 2·8 per 100 among boys and 2·5 per 100 among girls.

**Figure 2.**
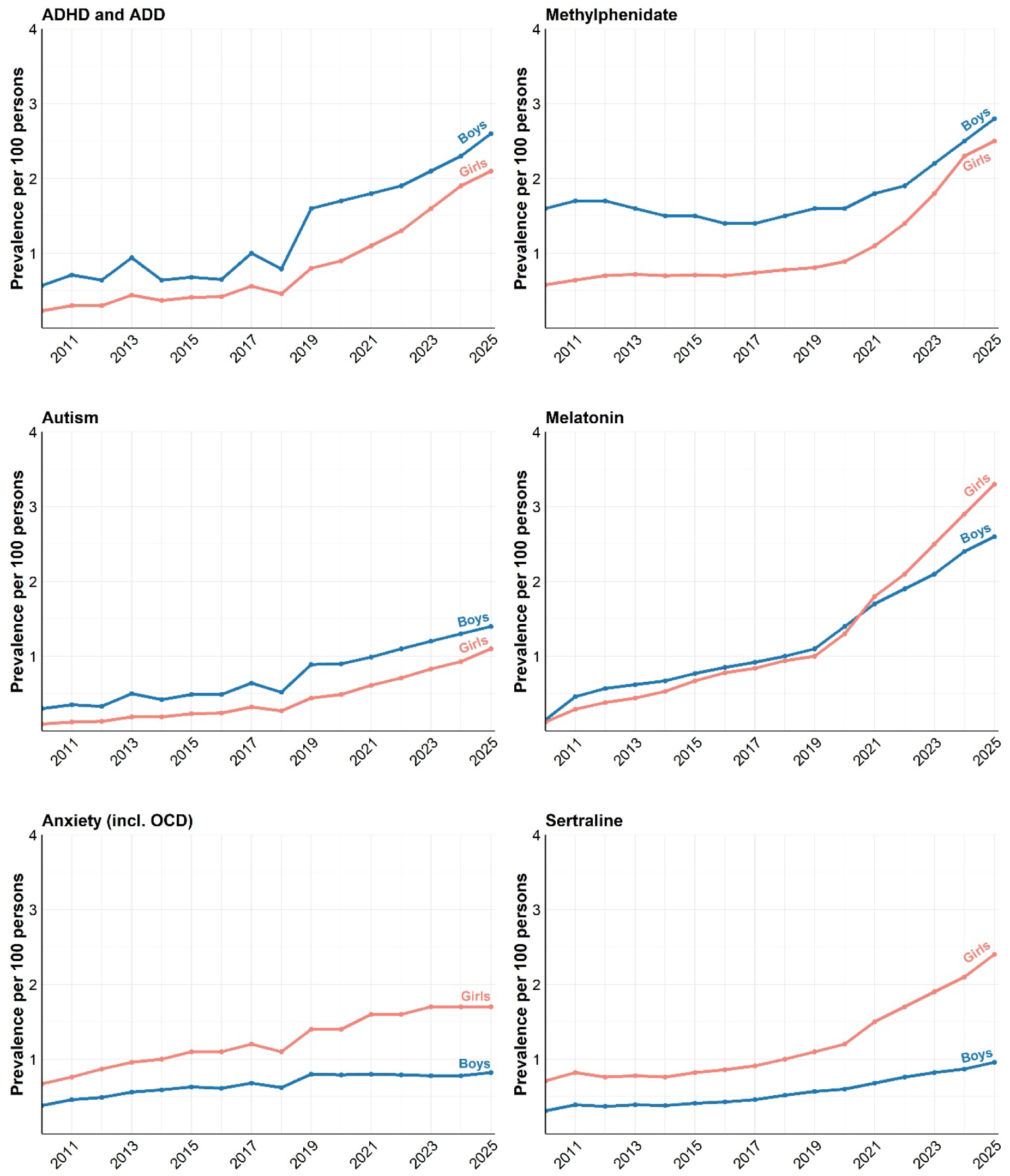
Sex-specific prevalence of selected psychiatric diagnoses and psychotropic medications. Annual prevalence per 100 persons for attention-deficit hyperactivity disorder (ADHD/ADD), autism spectrum disorders, anxiety, methylphenidate, melatonin, and sertraline among Danish residents aged 0–24 years from 2010 to 2025, stratified by sex assigned at birth. ADD: attention-deficit disorder; OCD: obsessive-compulsive disorder.

Age-stratified analyses showed that ADHD medication use increased steeply after 2020 for individuals aged six years and older and was highest among individuals aged 12–17, followed by individuals aged 18–24 years (**Figure 3**). Antidepressant use was most prevalent in the 18–24-year group, declining until 2016 and increasing thereafter. Melatonin use increased across all age groups throughout the study period, with a steeper rise around 2019, particularly among individuals aged 12–17 and 18–24 years.

**Figure 3.**
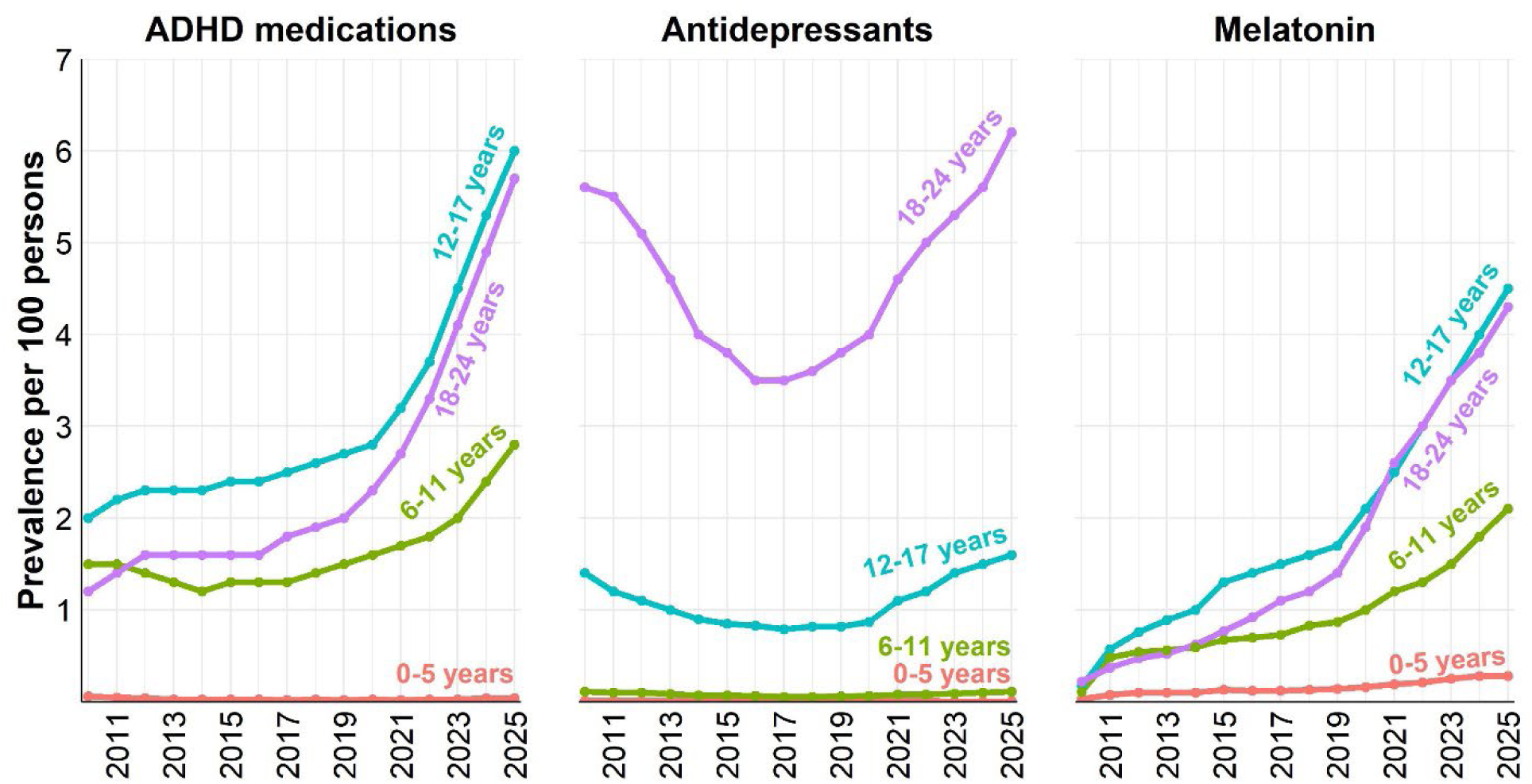
Age-stratified annual prevalence of selected psychotropic medication groups. Annual prevalence per 100 persons for ADHD medications (ATC code N06BA), antidepressants (N06A), and melatonin (N05CH01) among Danish residents aged 0–24 years from 2010 to 2025, stratified by age groups (0–5, 6–11, 12–17, and 18–24 years). Estimates are based on individuals alive and residing in Denmark on July 1^st^ of each year. ADHD: attention-deficit hyperactivity disorder

### Geographic variation

Geographic variation was evident at all examined levels. For the most-used medications across all regions in 2025, regional coefficients of variation ranged from 0·11 (sertraline) to 0·41 (atomoxetine) (**Figure 4**).

**Figure 4.**
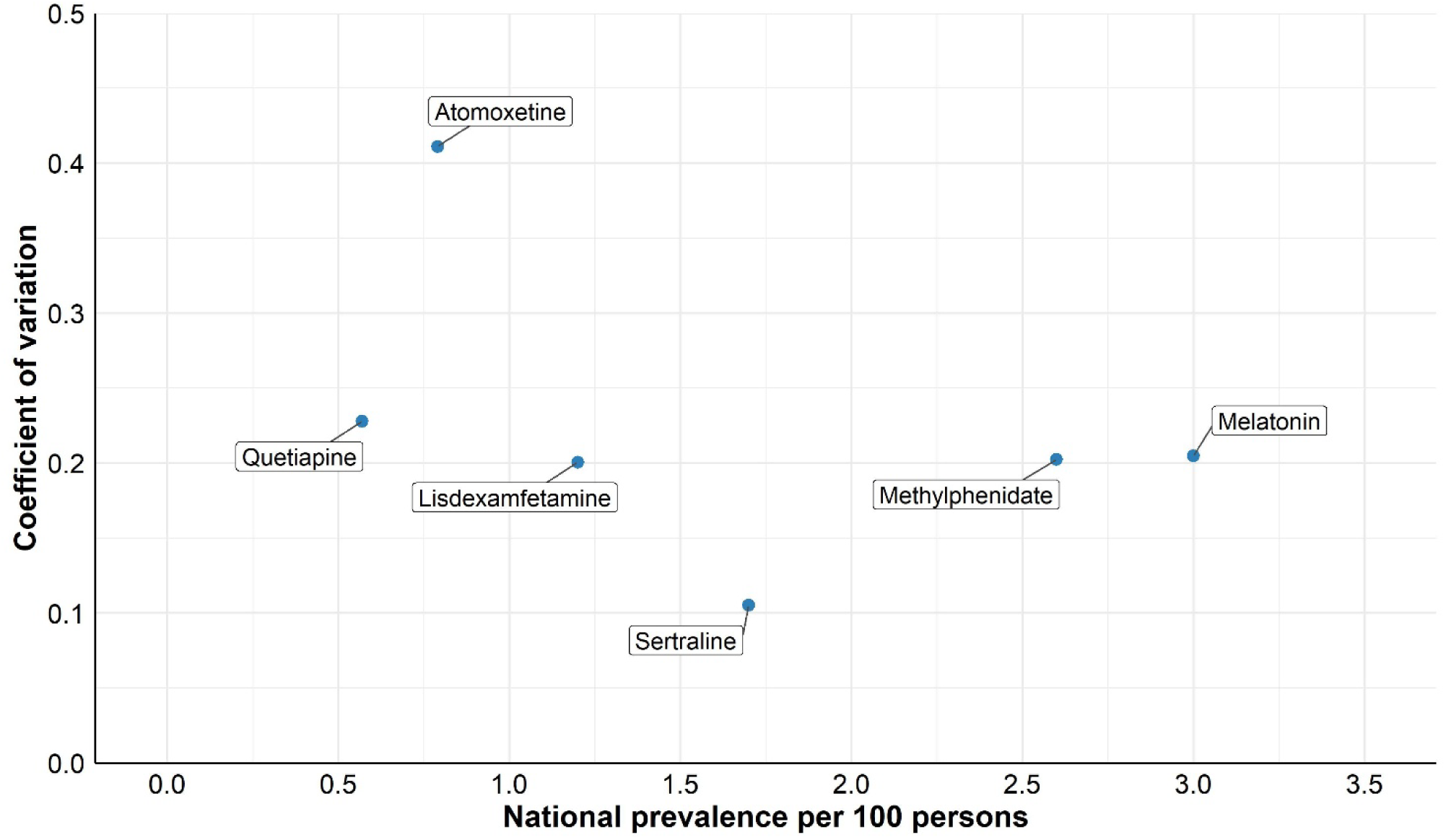
Geographic variation in prevalence of selected psychotropic medications across Danish regions in 2025. National prevalence per 100 persons plotted against the regional coefficient of variation for selected psychotropic medications across Danish regions in 2025 among residents aged 0–24 years. Each point represents a medication, and variation reflects differences across the five regions, calculated as the standard deviation divided by the national prevalence.

In 2025, atomoxetine use varied three-fold across health councils, ranging from 0·42 to 1·4 per 100 individuals aged 0–24 years (coefficient of variation 0·44) (**Figure 5**). Deviations from the national mean ranged from −47% in the lowest-rate areas to +77% in the highest-rate areas.

**Figure 5.**
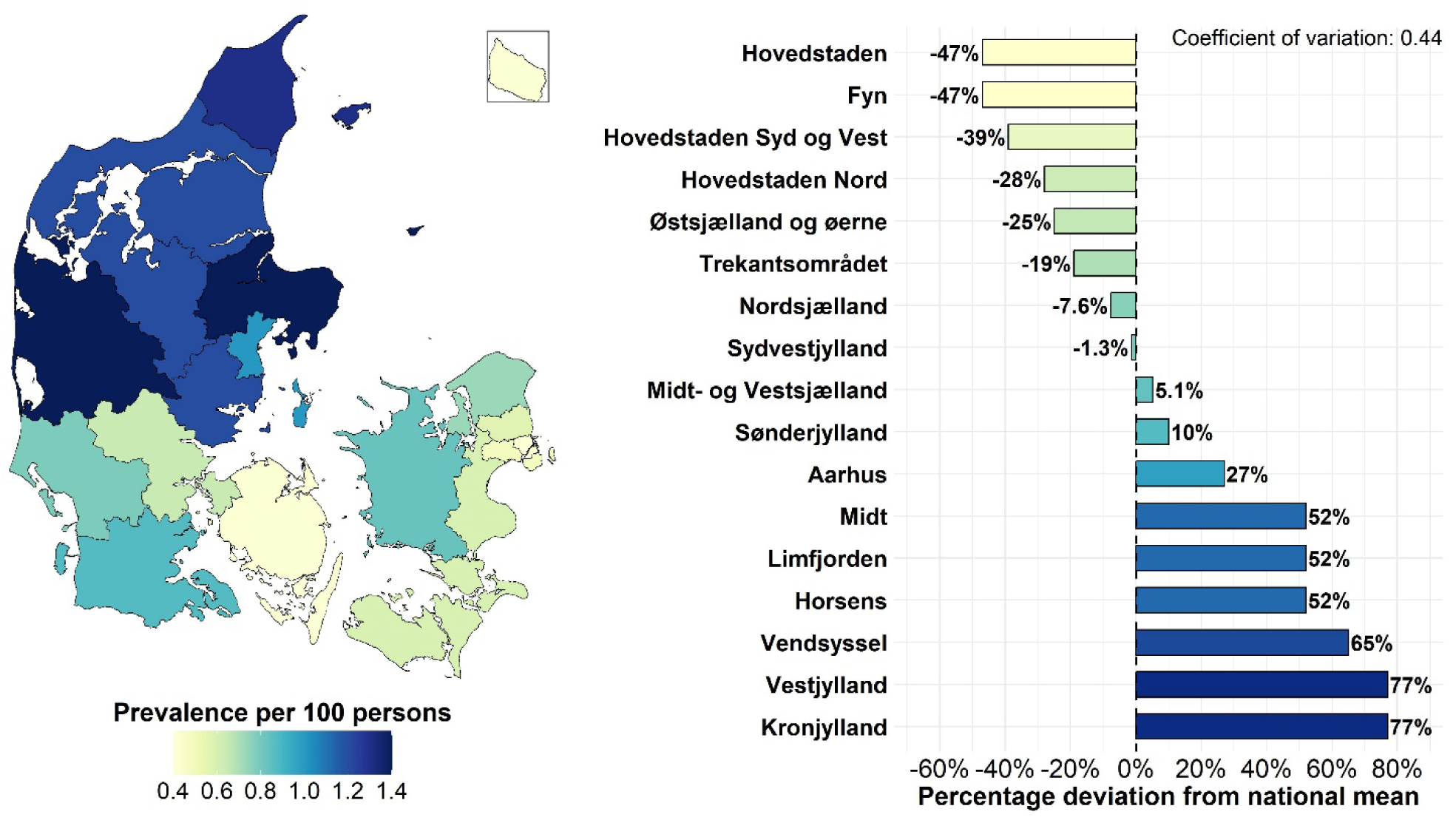
Prevalence of atomoxetine across Danish health councils in 2025. Age- and sex-standardised prevalence per 100 persons of atomoxetine across Danish health councils in 2025 among residents aged 0–24 years, and the percentage deviation from the national mean prevalence (0·79 per 100 persons) across health councils in 2025, with the coefficient of variation summarising geographic variation.

Geographic variation was also evident at the municipal level, as illustrated clearly by the lifetime prevalence of autism spectrum disorders in 2025. Lifetime prevalence was lowest in municipalities on Funen (1·3–2·2 per 100) and highest in parts of eastern Denmark (5·1–5·9 per 100; coefficient of variation 0·32) (**Figure 6**). The top 10% of municipalities accounted for 15% of the total prevalence across municipalities (Gini coefficient 0·17) (**Supplementary Figure 1**). Of the 97 municipalities with reportable prevalence proportions, 61 municipalities fell outside the 99·8% control limits in the funnel plot analysis (**Supplementary Figure 2**).

**Figure 6.**
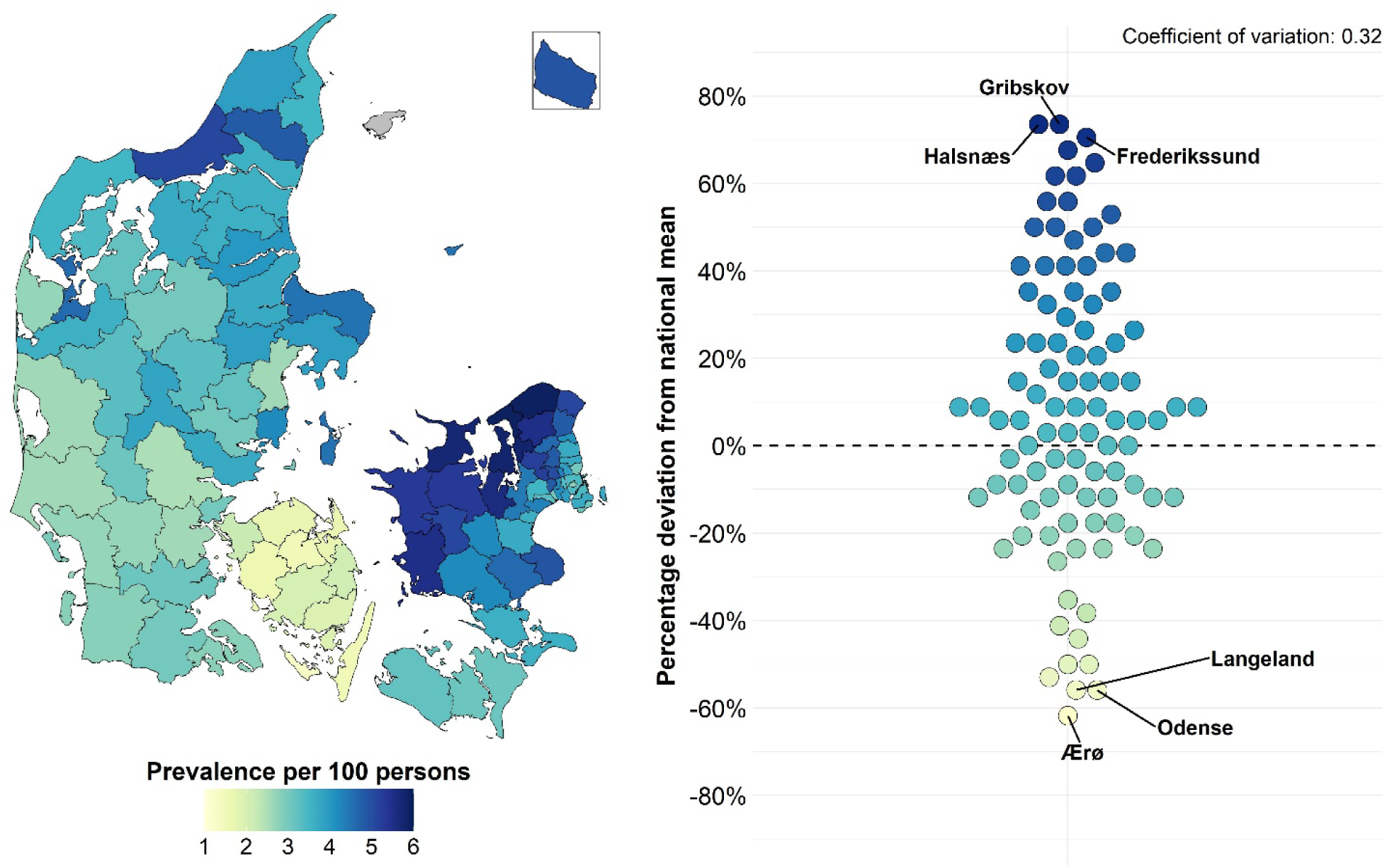
Municipality-level prevalence of autism in Denmark in 2025. Age- and sex-standardised prevalence per 100 persons of autism spectrum disorders across Danish municipalities in 2025 among residents aged 0–24 years, using the lifetime prevalence definition and percentage deviation in prevalence from the national mean (3·4 per 100 persons) across municipalities, with the coefficient of variation summarising geographic variation.

## Discussion

This nationwide study assessed psychiatric diagnoses and psychotropic medication use among Danish children, adolescents, and young adults from 2010 to 2025 and developed an interactive online tool to visualise temporal and geographic patterns. The findings showed substantial increases in psychiatric diagnoses and psychotropic medication use, particularly for ADHD, autism spectrum disorders, melatonin, methylphenidate, and sertraline. Geographic variation was evident across Danish regions, health councils, and municipalities. Although greater variation is expected in smaller population units, the magnitude and consistency of these differences suggest that factors other than underlying morbidity and random variation may contribute.

Key strengths include nationwide Danish health registers with near-complete population coverage, consistent application of diagnostic and medication definitions across years and geographic areas, and robust linkage through unique personal identifiers. Unlike studies based on insurance claims or selected samples, our data cover the entire population, so apparent variation is not driven by differences in coverage. Age and sex standardisation reduces the likelihood that observed geographic differences reflect variation in population composition. The study includes all redeemed prescriptions at community pharmacies and all hospital in- and outpatient contacts with a psychiatric disorder diagnosis, regardless of department, enabling consistent measurement over time and across geographic areas. Our framework and online tool can be applied in other countries with population-based registers.

Several limitations should be considered. First, we did not adjust for ethnicity, socioeconomic factors, underlying psychiatric morbidity, or patient preferences. Previous work indicates that socioeconomic position explains only a small share of geographic variation in ADHD and autism diagnoses.^6,7^ Still, as this is a descriptive study, our analyses cannot support causal inferences about the mechanisms underlying the variation. Second, diagnoses made outside the hospital sector are not captured in the Danish National Patient Registry,^20^ which may cause underestimation and bias geographic comparisons. However, in 2024, approximately 95% of diagnoses made by child and adolescent psychiatrists were registered by hospital-based services rather than private practice, with proportions ranging from 90% to 97% across the five regions.^25^ The corresponding proportions for other diagnosing professions could not be determined, so the overall extent of underestimation could not be quantified. In contrast, the prescription registry captures all prescriptions redeemed at community pharmacies regardless of prescriber, leaving out only in-hospital use.^21^ Third, geographic estimates at the municipal level are more sensitive to random fluctuation than estimates for larger administrative units. Although Danish municipalities are generally large enough for meaningful comparison, some small-area variation may reflect statistical instability rather than systematic differences, particularly in the least populated municipalities. However, for autism, 61 of 97 municipalities had prevalence estimates outside the 99·8% funnel plot control limits, indicating geographic heterogeneity beyond that expected under the statistical model. Such deviations do not establish whether differences reflect underlying differences in morbidity, diagnostic ascertainment, access to care, or clinical practice. Fourth, diagnostic validity and coding practices may differ between diagnostic categories, hospitals, and calendar periods, which may cause misclassification. Finally, temporal trends may partly reflect increased access to child and adolescent mental health services and increased demand for assessment and treatment over the study period.^26^ The findings may not be generalisable to health systems with a different organisation of child and adolescent mental healthcare or different reimbursement of psychotropic medication.

The rise in psychiatric diagnoses and psychotropic medication use among youths during the 16-year study period is consistent with previous Danish and international research. In particular, the marked growth in ADHD and autism diagnoses mirrors reported increases in cumulative incidence over time.^27,28^ Similarly, the upward trends in paediatric use of melatonin, methylphenidate, and sertraline align with findings from Denmark and other European settings, where increases have been documented across several classes of psychotropic medication.^2–4,29,30^

Geographic differences were evident across regions, health councils, and municipalities, in line with previous research. Similar differences in psychiatric diagnoses and psychotropic medication use among children and adolescents have been reported in Canada, the United Kingdom, and other countries,^5,30,31^ while municipal variation in ADHD and autism diagnoses has previously been documented in Denmark.^6,32^

The magnitude and consistency of these geographic differences suggest that variation in underlying morbidity, such as symptom burden or severity, may not fully explain them. International studies have found greater geographic variation in ADHD diagnosis rates than in symptom prevalence,^7^ and previous Danish research indicates that area-level characteristics, such as socioeconomic conditions, municipal spending, regional hospital waiting times, and the availability of private-practising child and adolescent psychiatrists, account for only part of the geographic differences.^32,33^ This suggests that differences in healthcare practices may play a central role, although our data cannot show the extent to which they contribute to the variation observed here.

At the healthcare-system level, differences in referral thresholds, local assessment capacity, and proximity to specialist care may influence if and when individuals receive a diagnosis or initiate treatment.^8^ Municipal differences in local educational and social services, including school-based support and psychological assessment services, may add to this.^34^ Clinician practices and local service cultures can also vary,^35^ and children from lower socioeconomic backgrounds are more frequently assigned behavioural diagnoses, potentially reflecting differences in disorder prevalence, help-seeking behaviour, or clinical assessment.^36^ Further research integrating individual-level, clinical, and service-level data is needed to explain the observed geographic patterns.

Our findings have practical implications for clinical practice and mental healthcare service planning. By providing nationwide estimates of temporal trends and geographic variation in psychiatric diagnoses and psychotropic medication use, together with an interactive online tool, this study enables identification of areas that deviate from national patterns. For example, the variation observed in atomoxetine use across health councils and in autism diagnoses at the municipal level may help prioritise areas for further clinical review and quality improvement efforts. Evidence from other healthcare systems indicates that reducing unwarranted variation requires more than descriptive data alone and is most effective when variation data are linked to local clinical insight and organisational decision-making.^11^ Our results provide a foundation for similar efforts in Denmark, including targeted quality-improvement initiatives, resource planning, and discussions of diagnostic and prescribing practices.

In conclusion, substantial increases in psychiatric diagnoses and psychotropic medication use, combined with geographic variation, were observed among Danish children, adolescents, and young adults between 2010 and 2025. These findings highlight differences in diagnostic and treatment patterns across geographic areas and underscore the importance of continued monitoring and evaluation of mental healthcare practice. The analytical approach used in this study, together with the interactive online tool, may support future efforts to examine variation and inform discussions of diagnostic and prescribing practices.

## Data Availability

The data used in this study are derived from Danish national health registers held by the Danish Health Data Authority. Because Danish legislation governs access to individual-level register data, the underlying data cannot be made publicly available. Researchers interested in accessing Danish register data may apply through the Danish Health Data Authority. The analysis code is available from the corresponding author on reasonable request. The study had no prespecified protocol and was not preregistered.

https://bupstat.sdu.dk/?lang=en

## Acknowledgements

We thank Peter Bjødstrup Jensen for validating the statistical analyses internally.

## Contributors

AP conceptualised and designed the study. HS conducted the statistical analyses, interpreted the results, and drafted the manuscript. HK, MB, RW, ASKH, AGO, and AP contributed to the study design and interpretation of results and critically revised the manuscript for important intellectual content. AP supervised the study. HS and AP directly accessed and verified the underlying data reported in the manuscript. The corresponding author (HS) had final responsibility for the decision to submit for publication.

## Supplementary Material

**Supplementary Table 1.** Characteristics of the study population in 2010 and 2025. Number and proportion of Danish residents aged 0–24 years alive on July 1^st^, 2010, and 2025, by sex assigned at birth, age group (0–5, 6–11, 12–17, and 18–24 years) and residential region. Counts correspond to the denominator used in the prevalence analyses. We included 2,930,744 unique individuals across 2010–2025.

| Characteristics | 2010<br>N = 1,675,958 | 2025<br>N = 1,651,616 |
| --- | --- | --- |
| **Sex, n (%)** |  |  |
| Male | 858,414 (51) | 846,714 (51) |
| Female | 817,544 (49) | 804,902 (49) |
| **Age, median (IQR)** | 12 [6-18] | 13 [6-19] |
| **Age group, n (%)** |  |  |
| 0–5 years | 391,506 (23) | 364,532 (22) |
| 6–11 years | 397,953 (24) | 373,201 (23) |
| 12–17 years | 422,770 (25) | 399,359 (24) |
| 18–24 years | 463,729 (28) | 514,524 (31) |
| **Residential region, n (%)** |  |  |
| Capital Region of Denmark | 499,675 (30) | 529,939 (32) |
| Zealand Region | 239,167 (14) | 215,624 (13) |
| Region of Southern Denmark | 363,744 (22) | 328,817 (20) |
| Central Denmark Region | 398,117 (24) | 389,311 (24) |
| North Denmark Region | 173,968 (10) | 156,695 (9·5) |
| Unknown | 1,287 (< 0·1) | 31,230 (1·9) |

IQR: interquartile range.

**Supplementary Figure 1.**
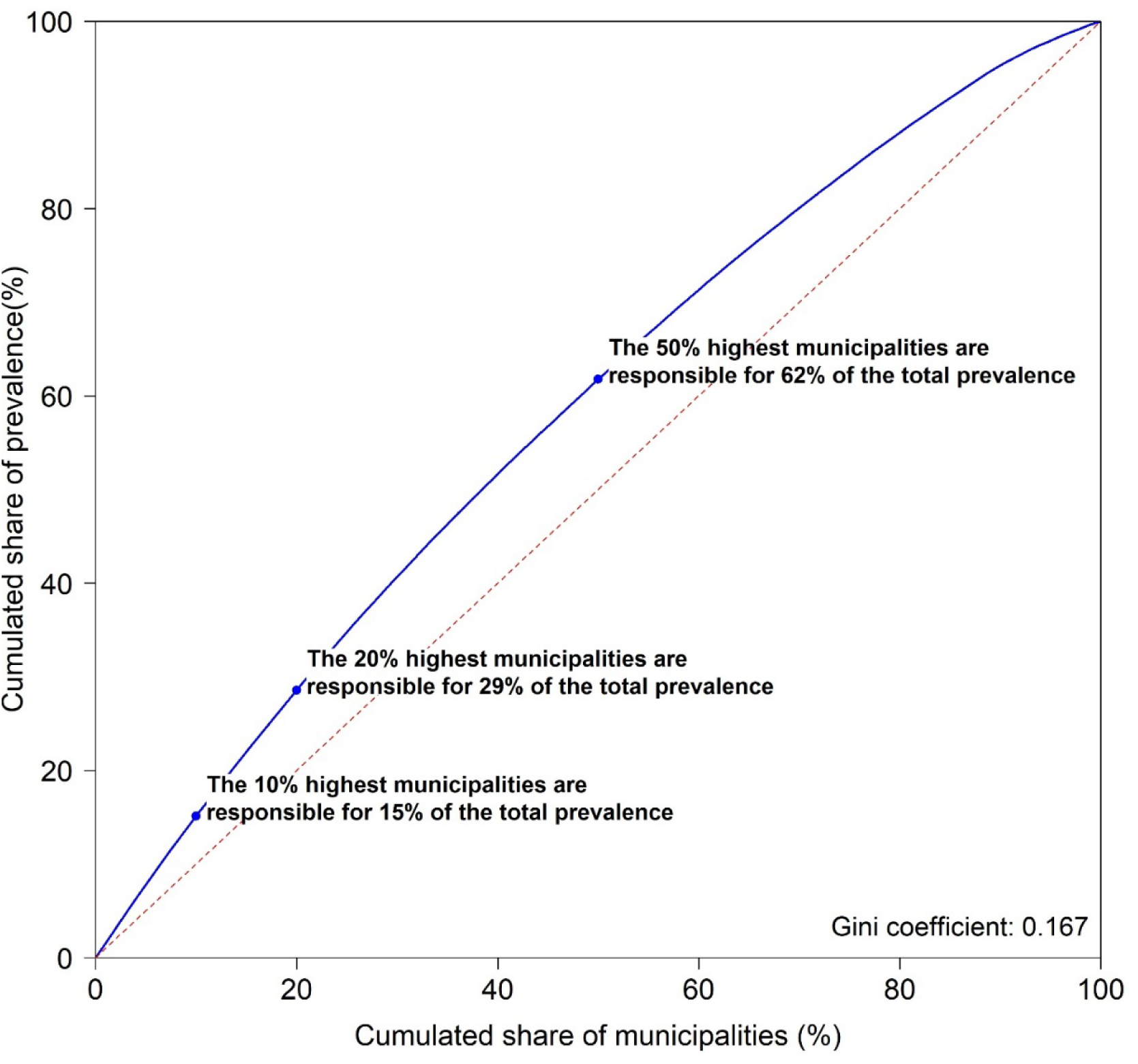
Lorenz curve for municipality-level inequality in autism prevalence in Denmark among residents aged 0–24 years in 2025. The Lorenz curve shows the cumulative share of autism lifetime prevalence accounted for by the cumulative share of municipalities ranked by prevalence among residents aged 0–24 years in 2025. The corresponding Gini coefficient summarises inequality.

**Supplementary Figure 2.**
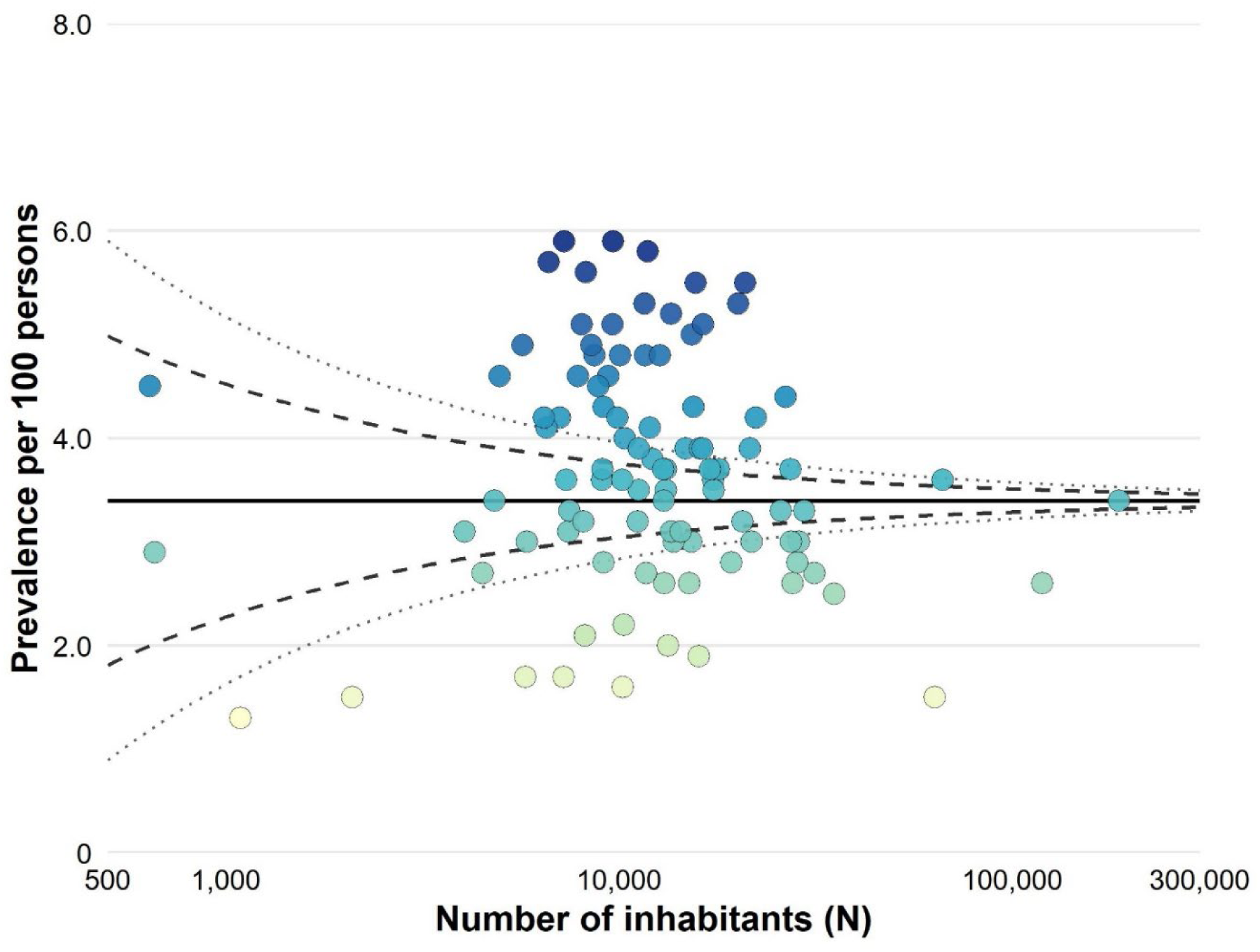
Funnel plot of municipality-level variation in autism prevalence in Denmark among residents aged 0–24 years in 2025. Each point represents a municipality, plotted as lifetime prevalence of autism spectrum disorders per 100 persons against the number of inhabitants aged 0–24 years (logarithmic scale). The solid horizontal line shows the national mean (3·4 per 100 persons). The dashed and dotted curves show the 95% and 99·8% control limits, respectively, which indicate the expected range of variation due to random fluctuation around the national mean under a binomial distribution. Point colour corresponds to prevalence.

## Appendix

**Appendix Table 1.** Definition of psychiatric diagnoses and psychotropic medications according to the International Classification of Diseases, 10th Revision (ICD-10) and ATC codes.

| **Psychiatric diagnosis** | **ICD-10 codes** | **Incidence definition** | **Annual prevalence definition** | **Lifetime prevalence definition** |
| --- | --- | --- | --- | --- |
| Schizophrenia spectrum disorder | F20-29 | First diagnosis ever occurring in the current year | A diagnosis in the current year | A diagnosis in the current year or earlier years |
| Bipolar disorder | F30-31 | First diagnosis ever occurring in the current year | A diagnosis in the current year | Not estimated |
| Depressive episode | F32-33 | First diagnosis ever occurring in the current year | A diagnosis in the current year | Not estimated |
| Anxiety disorders incl. obsessive-compulsive disorder | F40-48 plus F93 | First diagnosis ever occurring in the current year | A diagnosis in the current year | Not estimated |
| Eating disorder | F50 | First diagnosis ever occurring in the current year | A diagnosis in the current year | Not estimated |
| Personality disorder | F60-F69 | First diagnosis ever occurring in the current year | A diagnosis in the current year | A diagnosis in the current year or earlier years |
| Intellectual disability | F70–79 | First diagnosis ever occurring in the current year | A diagnosis in the current year | A diagnosis in the current year or earlier years |
| Autism spectrum disorder | F84 excluding F84.2-F84.4 | First diagnosis ever occurring in the current year | A diagnosis in the current year | A diagnosis in the current year or earlier years |
| Attention-deficit hyperactivity disorder and attention-deficit disorder | F90, F98.8 | First diagnosis ever occurring in the current year | A diagnosis in the current year | A diagnosis in the current year or earlier years |
| Attachment disorder of childhood | F94 excluding F94.0 | First diagnosis ever occurring in the current year | A diagnosis in the current year | A diagnosis in the current year or earlier years |
| Tic disorders | F95 | First diagnosis ever occurring in the current year | A diagnosis in the current year | Not estimated |
| **Psychotropic medication** | **ATC codes** | **Incidence definition** | **Prevalence definition** |  |
| Psycholeptics | N05* | First redemption ever occurring in the current year | A redemption of a prescription in the current year |  |
| Psychoanaleptics | N06* | First redemption ever occurring in the current year | A redemption of a prescription in the current year |  |

\* Analyses were conducted across all four ATC hierarchy levels
ATC: Anatomical Therapeutic Chemical; ICD-10: International Classification of Diseases, 10th Revision.

**Appendix Table 2.** Specific ATC codes for psychotropic medications mentioned in the manuscript.

| **Psychotropic medication** | **ATC code** | **Group of medication** |
| --- | --- | --- |
| Quetiapine | N05AH04 | Diazepines, oxazepines, thiazepines and oxepines |
| Melatonin | N05CH01 | Melatonin receptor agonists |
| Sertraline | N06AB06 | Selective serotonin reuptake inhibitors |
| Duloxetine | N06AX21 | Other antidepressants |
| Methylphenidate | N06BA04 | Centrally acting sympathomimetics |
| Atomoxetine | N06BA09 | Centrally acting sympathomimetics |
| Lisdexamfetamine | N06BA12 | Centrally acting sympathomimetics |

ATC: Anatomical Therapeutic Chemical.

**Appendix Table 3.**
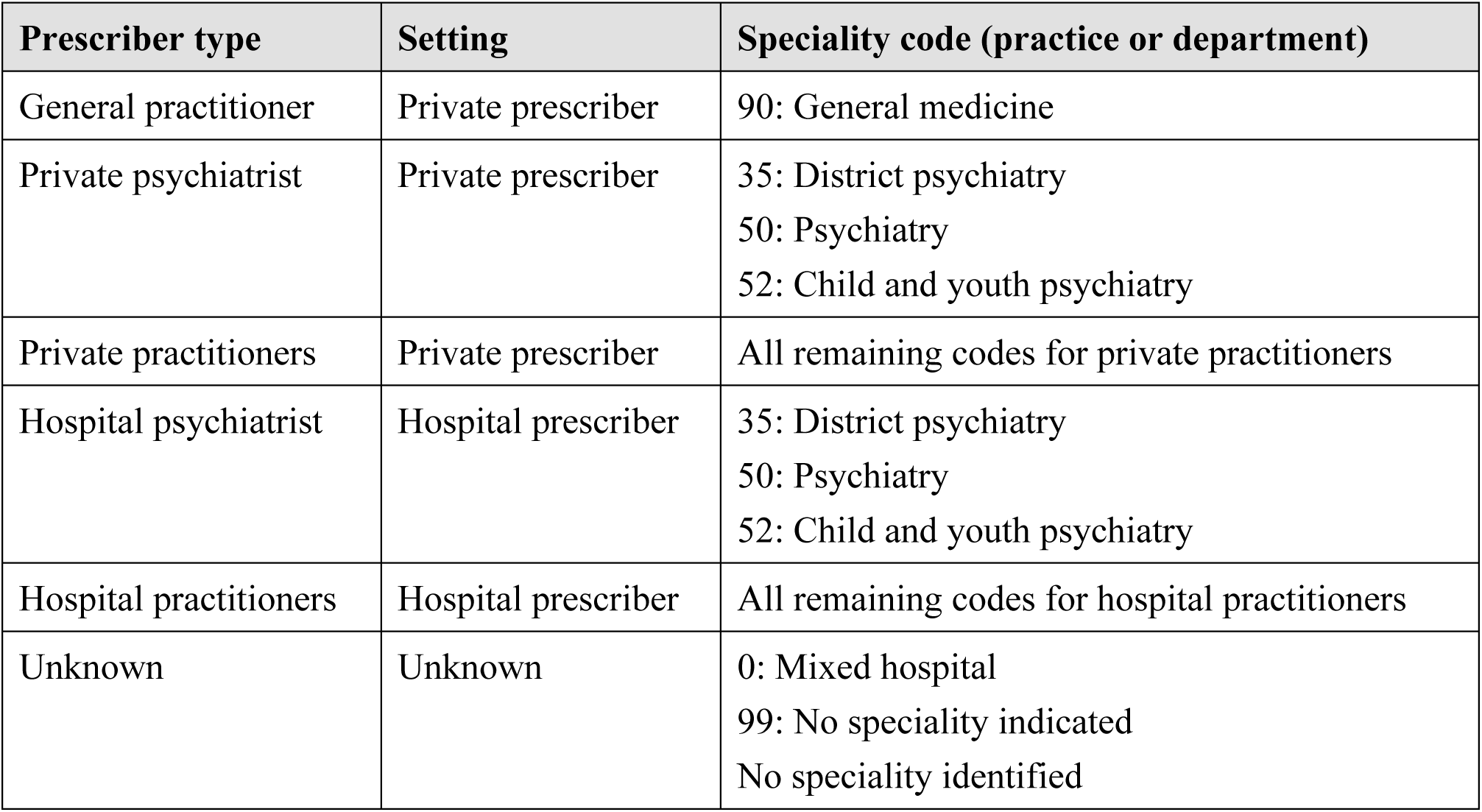
Definition of the different types of prescribers with codes obtained from the Registry of Health Care Providers.

| **Prescriber type** | **Setting** | **Speciality code (practice or department)** |
| --- | --- | --- |
| General practitioner | Private prescriber | 90: General medicine |
| Private psychiatrist | Private prescriber | 35: District psychiatry<br>50: Psychiatry<br>52: Child and youth psychiatry |
| Private practitioners | Private prescriber | All remaining codes for private practitioners |
| Hospital psychiatrist | Hospital prescriber | 35: District psychiatry<br>50: Psychiatry<br>52: Child and youth psychiatry |
| Hospital practitioners | Hospital prescriber | All remaining codes for hospital practitioners |
| Unknown | Unknown | 0: Mixed hospital<br>99: No speciality indicated<br>No speciality identified |

### Appendix Text 1 PubMed search strings. Searches were run on 25 September 2026 with no date or language filters.

Search 1, geographic variation (1,257 records):

(“psychiatric disorder*”[tiab] OR “mental disorder*”[tiab] OR “mental health”[tiab] OR ADHD[tiab] OR “attention deficit”[tiab] OR autis*[tiab] OR psychotropic*[tiab] OR antidepressant*[tiab] OR methylphenidate[tiab] OR melatonin[tiab]) AND (child*[tiab] OR adolescen*[tiab] OR “young adult*”[tiab] OR youth[tiab]) AND (geograph*[tiab] OR region*[tiab] OR municipal*[tiab] OR “small area”[tiab] OR atlas[tiab]) AND (variation[tiab] OR variability[tiab] OR trend*[tiab])

Search 2, time trends in Denmark and other Nordic countries (297 records):

(“psychiatric disorder*”[tiab] OR “mental disorder*”[tiab] OR ADHD[tiab] OR “attention deficit”[tiab] OR autis*[tiab] OR psychotropic*[tiab] OR antidepressant*[tiab] OR methylphenidate[tiab] OR melatonin[tiab]) AND (child*[tiab] OR adolescen*[tiab] OR “young adult*”[tiab]) AND (trend*[tiab] OR incidence[tiab] OR prevalence[tiab] OR utilisation[tiab] OR utilization[tiab]) AND (Denmark[tiab] OR Danish[tiab] OR Nordic[tiab] OR Scandinavia*[tiab])

